# A phased lift of control: a practical strategy to achieve herd immunity against Covid-19 at the country level

**DOI:** 10.1101/2020.03.29.20046011

**Authors:** Sake J. de Vlas, Luc E. Coffeng

## Abstract

Most countries are affected by the Covid-19 pandemic and experience rapidly increasing numbers of cases and deaths. Many have implemented nationwide stringent control to avoid overburdening the health care system. This paralyzes economic and social activities until the availability of a vaccine, which may take years. We propose an alternative exit strategy to develop herd immunity in a predictable and controllable way: a phased lift of control. This means that successive parts of the country (e.g. provinces) stop stringent control, and Covid-19-related IC admissions are distributed over the country as the whole. Importantly, vulnerable individuals need to be shielded until herd immunity has developed in their area. We explore the characteristics and duration of this strategy using a novel individual-based model for geographically stratified transmission of Covid-19 in a country. The model predicts that individuals will have to experience stringent control for about 14 months on average, but this duration may be significantly shortened by future developments (more IC beds, better drugs). Clearly, the strategy will have a profound impact on individuals and society, and should therefore be considered carefully by various other disciplines (e.g. health systems, ethics, economics) before actual implementation.

---

Most parts of the world are affected by the pandemic of coronavirus disease (Covid-19) and experience rapidly increasing numbers of cases and deaths, with fatalities mainly occurring among the old and otherwise vulnerable. Some countries (e.g. Italy) are experiencing an overload of patients requiring intensive care (IC) facilities, leading to heart-breaking triage decisions.^1^ Most countries have implemented nationwide stringent control efforts, which paralyze economic and social activities. Some (e.g. France and Belgium) have even imposed a lock-down.

China and South Korea have demonstrated that with very intensive interventions viral transmission can be pushed down to low levels,^2^ but this will not offer a permanent solution in the foreseeable future. Without sufficient herd immunity, the Covid-19 epidemic will revert to its original dynamic course as soon as interventions are withdrawn.^3^ This resurgence can only be prevented if most countries in the world follow China’s example and jointly maintain intensive control for a long time, probably years, until the very last cases have been tracked down and isolated. Furthermore, countries with residual circulating virus would have to be completely isolated to avoid re-introduction. Still, prolonged intensive control could be a means to gain time until the development and large-scale availability of a vaccine. However, the amount of time this will take is unpredictable. An alternative approach is to develop herd immunity through natural infection while keeping the number of cases within the limits of the health care system. Until recently, the UK has advocated this approach,^4^ but it was heavily criticized for obvious reasons: it will be practically impossible to perfectly tune actual interventions without exceeding or undershooting health care capacity. Clearly, there is a need for an exit strategy that is predictable and controllable. We believe that we have found such a strategy.

We propose that countries consider “a phased lift of control”. That is, in successive parts of the country (say provinces or municipal health services catchment areas) all stringent interventions are released, such that the epidemic can rage locally, while maintaining strict control in the remaining parts that wait for their turn. At the same time, Covid-19-related IC admissions should be distributed over the whole country such that the national health care system is not overburdened. After the lift of control in their area, its inhabitants can resume their normal daily activities as before Covid-19. Importantly, individuals most vulnerable to the virus need to be shielded until their area has achieved herd immunity. A further requirement is that their care providers (professionals or possibly family members) are absolutely free of the virus or immune. Obviously, this strategy will require a lot of ambulance transport of severe cases, but this should be feasible in small countries with good infrastructure, such as the Netherlands. Larger countries can consider implementing the strategy in specific geographic regions. Box 1 provides a summary of the strategy, including minimum conditions and a calculation of the IC capacity threshold in terms of the number of prevalent Covid-19 cases in the population.

## Box 1 A phased lift of control to achieve herd immunity against Covid-19

### The strategy

A large geographic area (say country) is subdivided into parts (e.g. provinces). Control is lifted in one part, so that a Covid-19 epidemic ensues locally. To avoid overburdening of the health care system, all Covid-19-related IC admissions are distributed over the whole country. Once case numbers peak, control is lifted in the next part. This is repeated until all parts of the country have lifted control.

### Minimum conditions

- Shielding of vulnerable cases until their local area has achieved herd immunity
- Geographic area has sufficient infrastructure to transport IC patients between all parts
- Control is sufficiently intensive to avoid outbreaks in the parts that have not yet lifted control

### Determinants of IC capacity threshold

1. Number of IC beds available for Covid-19 patients in the entire country
2. Fraction of Covid-19 cases eventually requiring IC admission
3. The average duration of IC admission

All three may improve in the near future, allowing parts of the country to lift control earlier, thereby reducing the overall duration of the strategy. See below for values for the Netherlands.

### Illustration (based on Figure 1)

Model-predicted number of infectious Covid-19 cases per million during a phased lift of control in 10 successive areas (colored lines), with a first interval of 120 days and 90 days from thereon (vertical dashed lines), after an initial phase of 15 days of nation-wide intensive control. The overall number of cases (solid black line) remains well below the threshold associated with the maximum IC capacity.

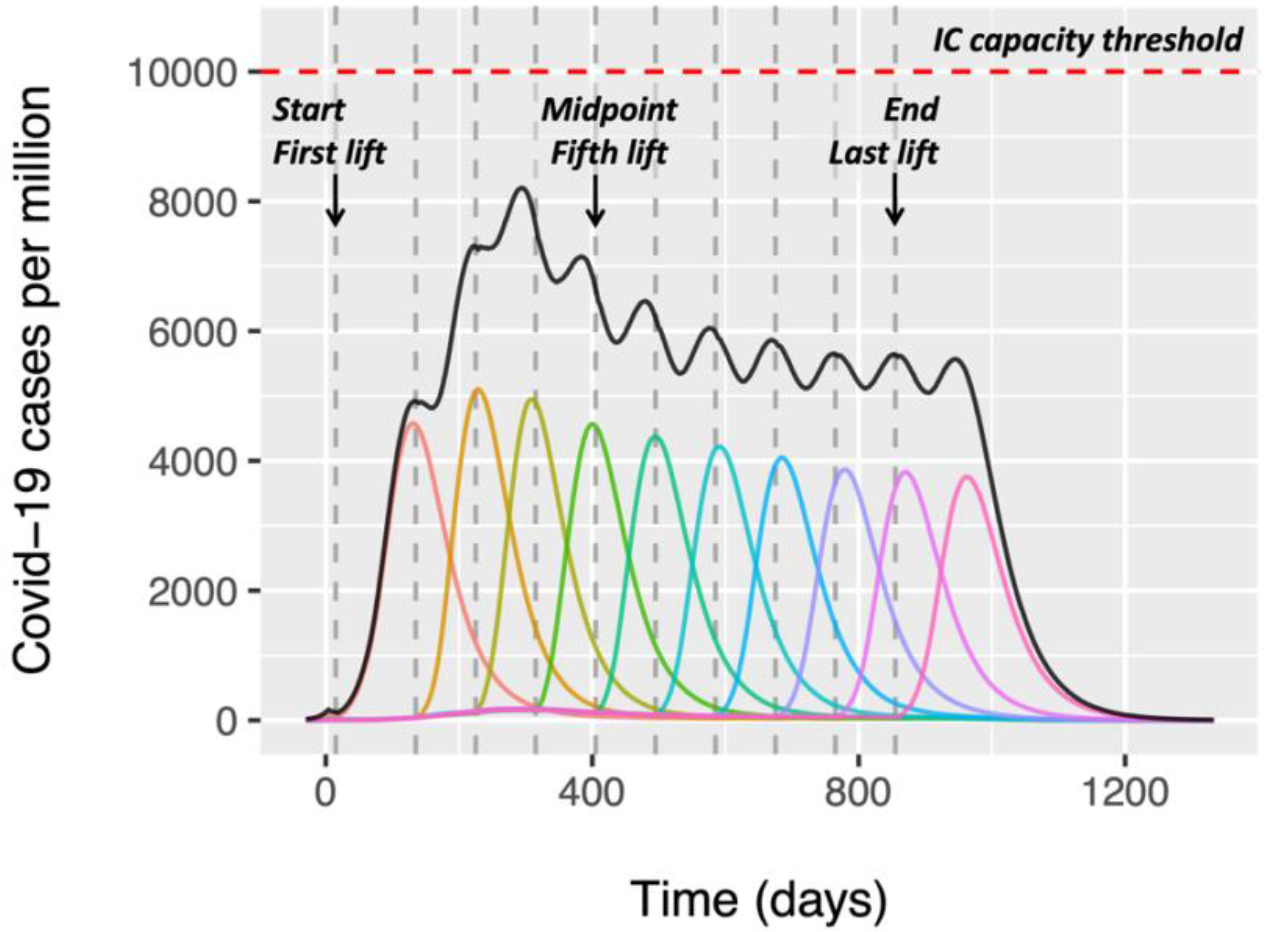

### IC capacity threshold for the Netherlands

First, normally the Netherlands (17.4 million inhabitants) has 1150 IC beds, of which half (575) would be available for Covid-19 cases. Recently, this capacity has been increased to 1500 IC beds, and this will be extended to 2,400 IC beds, of which still 500 are needed for other conditions. Overall, this amounts to a maximum IC capacity of about 1,900 IC beds for patients with severe Covid-19 morbidity, i.e. 109.2 IC beds per million. Second, of all known Dutch Covid-19 cases, 1 in 4 requires hospital care, of whom 1 in 4 needs IC (1/16 in total). The actual number of Covid-19 cases in the Netherlands is unknown, but may be estimated using data from Germany where intensive contact tracing and testing has been performed in the general population. As of 2 April 2020, Germany had 73,552 recognized Covid-19 cases of whom 827 died ([Robert Koch Institute](https://www.rki.de/DE/Content/InfAZ/N/Neuartiges_Coronavirus/Situationsberichte/2020-04-02-de.pdf?__blob=publicationFile)), and by 4 April the Netherlands reported 16,627 confirmed cases and 1,651 fatalities ([RIVM](https://www.rivm.nl/coronavirus-covid-19/actueel)). Assuming that actual case fatality rates in both countries are similar, there are 8.83 Covid-19 cases (1,651/16,627 ÷ 827/73,552) for every recognized case in the Netherlands. Thus, 1 in 141.3 (16 × 8.83) of all Covid-19 cases eventually requires IC admission. Third, Covid-19-related IC admissions typically last about 3 to 4 weeks (say 24 days), which is 2.4 times the presumed duration that an individual is infectious/ symptomatic (on average 10 days). This leads to an IC capacity threshold in terms of the prevalent number of Covid-19 cases equal to 109.2 × 141.3 × 10/24 = 6429 per million. As Germany probably did not identify all Covid-19 cases, we expect that there are even more than 8.83 Covid-19 cases for every recognized case in the Netherland. We therefore round the threshold up to 10 thousand per million.

### Illustration of an optimistic scenario

Model-predicted number of infectious Covid-19 cases per million during a phased lift of control in 10 successive areas (colored lines), with a 90-90-60-45-45-30-30-30-30 days scheme of intervals (vertical dashed lines), after an initial phase of 15 days of nation-wide intensive control. The overall number of cases (solid black line) remains below the threshold associated with the maximum IC capacity, anticipating a further doubling of capacity over time, e.g. due to more IC beds, or new drugs and better shielding of risk populations leading to lower IC need and/or shorter duration of IC admission.

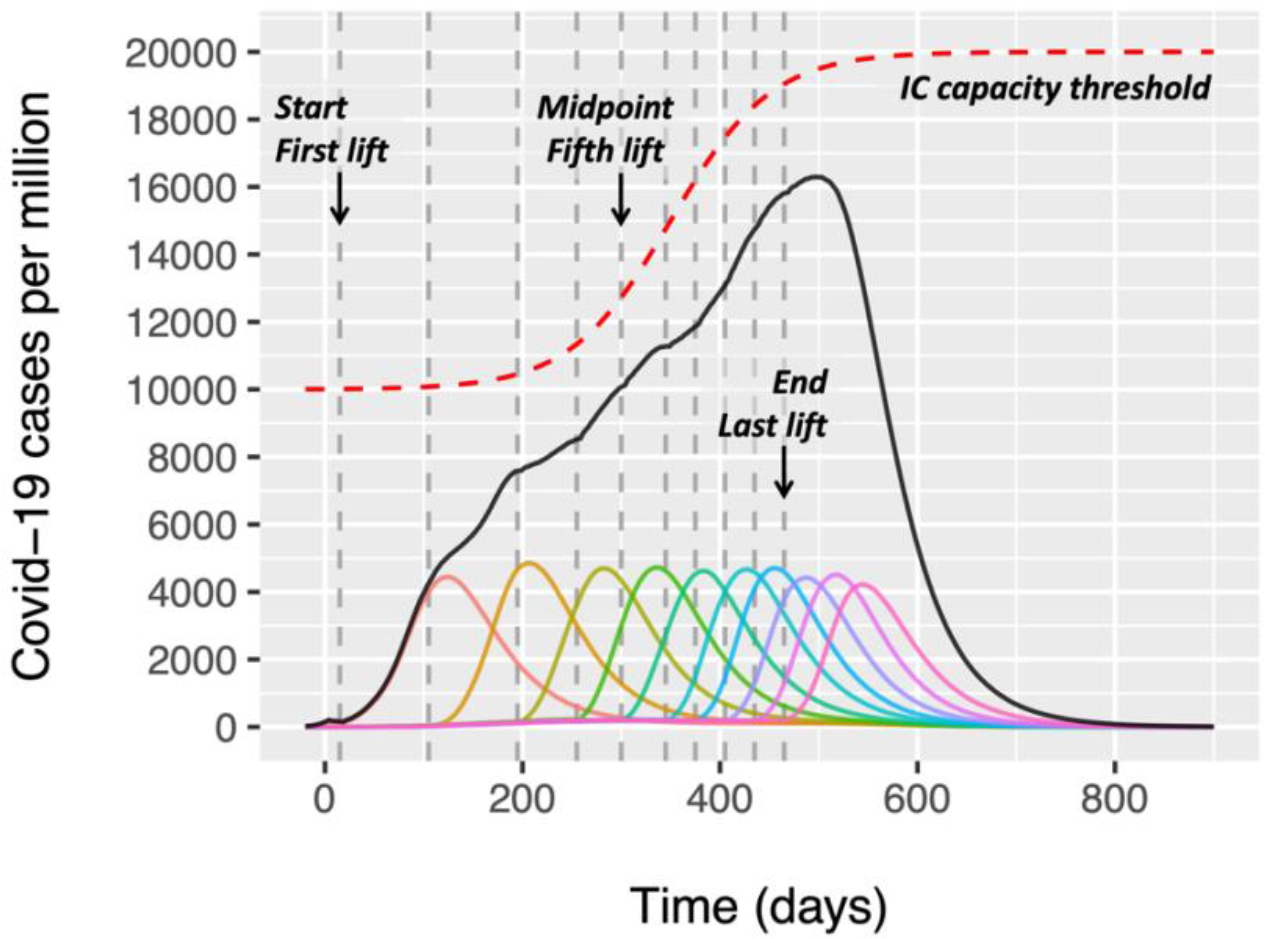

To explain the potential impact, let’s do a back-of-the-envelope calculation taking the Netherlands context as an example. The country can harbor at most 10 thousand prevalent infectious cases per million population without overstretching the health care system (Box 1). At this maximum, the average flow (incidence) is about 1000 new Covid-19 cases per day, i.e. 10 thousand prevalent cases divided by the presumed average duration of infectiousness of 10 days.^3^ Given the estimated basic reproduction number *R*_0_ = 2.5,^3,5^ at least 60% (1 – 1/*R*_0_) of the population should have experienced the infection and have acquired immunity in order to provide herd immunity to the remaining 40%. To achieve the required 600 thousand immune individuals per million population – under optimal use of healthcare capacity – will take at least 600 days (600 thousand/1000 new cases per day). Now suppose we implement the strategy of a phased lift of control and divide a hypothetical country with 1 million inhabitants into 10 equal parts (say provinces) with 100 thousand inhabitants each. We then lift control in the first selected province and allow, e.g., 75% of the national supply of IC beds to be used for Covid-19 patients from this province. The remaining 25% can be used for severe Covid-19 cases in the rest of the country. Then the selected province can harbor 7500 cases (i.e. 7.5 times as many as a strategy focusing on the country as a whole). Thus, ideally, only 600/7.5 = 80 days are needed to reach the required number of immunes in this province. Subsequently, the next province can be chosen to lift control, again requiring about 80 days, and so on. The 10-th and last province will lift control on day 720. This is somewhat longer than the 600 days above, but on average individuals in the country will be free of interventions after 360 days.

There are several factors that influence the duration of this strategy in practice. First of all, local outbreaks will not show stable levels but typically display epidemic peaks, effectively requiring the average number of cases to stay well below the IC threshold. This will obviously prolong the overall duration needed. On the other hand, some efficiency will be gained from heterogeneity in contact rates: initially those with many daily contacts will acquire the infection first, such that those who remain uninfected (non-immune) in the end phase tend to be those with fewer contacts. This will decrease the minimum required level of herd immunity. Furthermore, time could be saved by starting the moments of lifting control earlier, such that local epidemics somewhat overlap. Understanding the complex balance between these three processes (peaky behavior, selection of those with fewer contacts, and partial overlap of local epidemics) requires mathematical modelling.

We have therefore developed a stochastic individual-based model to explore how this strategy of a phased lift of control may turn out in practice. For the example in Figure 1, the model simulates a population of 10 million individuals, living in 10 thousand clusters (towns, wards or villages) that are grouped in 10 about equally sized superclusters (e.g. provinces). The clusters vary in size such that the 95% extremes (i.e. 2.5% and 97.5% percentiles) reflect populations of about 100 to 4000 inhabitants, with the largest 0.2% exceeding 10 thousand. Heterogeneity is introduced by varying individual contact rates (i.e. the combination of contact frequency and transmission probability) according to a distribution such that both 95% extremes represent a 10-fold relative difference. Furthermore, there is some degree of assortative mixing by allowing mean contact rates to vary more than randomly among clusters. As a consequence, the average contact rate is 4 times higher for the 97.5% vs. 2.5% percentile of all clusters. We describe Covid-19 transmission as a standard SEIR (susceptible, exposed, infectious/symptomatic, recovered/immune) process, assuming life-long immunity. Durations of the exposed and infectious stages are assumed to be 5.5 and 10 days, respectively, similar to a recent modelling exercise.^3^ The model further captures three levels of transmission: within clusters (90% weight), within superclusters (5%), and within the entire population (5%). The start of the epidemic is simulated by randomly seeding 10 infections in one supercluster. The average contact rate has been tuned such that the initial exponential increase of Covid-19 cases matched that of a fully homogeneous SEIR model with *R*_0_ = 2.5. Intensive control is assumed to start when the epidemic reached a cumulative number of 300 cases per million. Intensive control is modelled as a reduction of the average contact rate, which we have chosen to be 25% of its original value (75% reduction). Lifting control within a supercluster is assumed to lead to contact rates immediately returning to and staying at their original values for the remainder of the simulation. We further assume some degree of isolation of the supercluster that just lifted control by halving its contribution and exposure to transmission at the overall population level. The chosen critical threshold is 10 thousand prevalent infectious Covid-19 cases per million population, corresponding to the above assumed maximum number of IC beds in the Netherlands. Supplement 1 provides a full technical description of the model.

**Figure 1.**
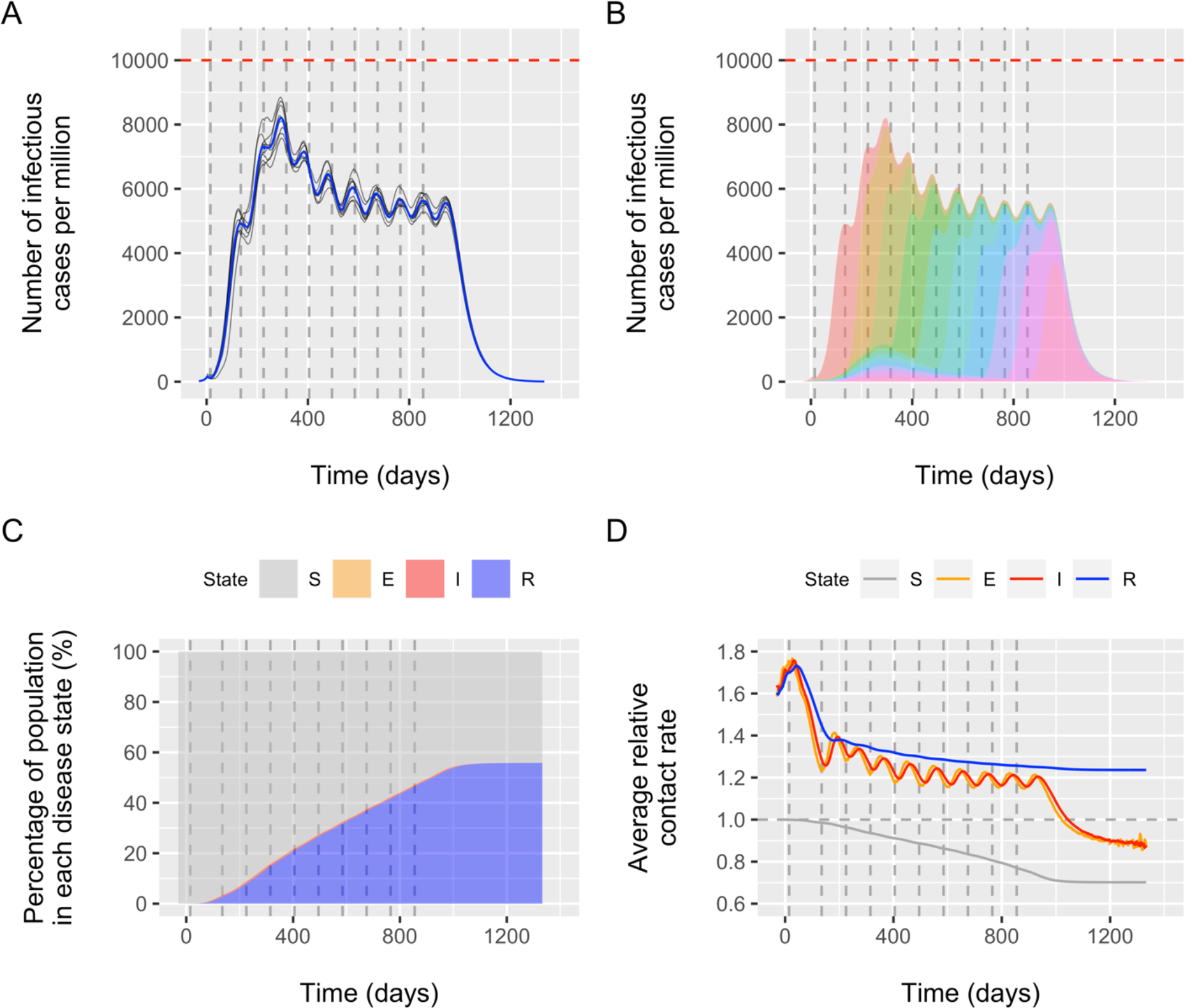
Model-predicted outcome of a phased lift of control against Covid-19 in a population of 17 million (the Netherlands). The population (a country or part of a country) is divided over 10 superclusters (provinces or municipal health services catchment areas), each harboring 1700 clusters (towns, wards or villages) with on average 1000 inhabitants. Panel A: overall number of Covid-19 cases per 1 million for 8 model runs (average trend in blue). Panel B: contribution of each of the 10 superclusters (colored areas) to the overall average number of cases. Panel C: proportion of the population in the modelled disease states: susceptible (S), exposed (E), infectious/symptomatic (I), and removed/immune (R). Panel D: average relative contact rate of individuals in each disease state. Time is defined in terms of days since onset of intensive control in the entire population. The 10 vertical dashed lines indicate the timing of the moments of (permanently) lifting control in the successive superclusters. In the first supercluster, the one with the highest initial burden, lifting of control occurred after 15 days, a (random) second supercluster followed 120 days later, followed by a phased lifting of control in another supercluster each 90 days (random order). The horizontal red line in (A) and (B) indicates the threshold of 10 thousand Covid-19 cases per million, which corresponds to the maximum IC capacity in the Netherlands (Box 1).

Figure 1A shows that with a strategy of a phased lift of control every 90 days across 10 superclusters, the average trend in the overall number of Covid-19 cases remains well under 10 thousand per million. This interval of 90 days is longer than the 80 days used in the example above to account for the peaky behavior. Control was lifted first in the most affected supercluster (i.e. Noord-Brabant province in the Netherlands), as this provided the least risk of exceeding the threshold; the order of subsequent superclusters was random. Further, the interval between lifting control in the first and second supercluster was allowed to be longer (here we chose 120 days) to let the first local outbreak to reach its peak. The fluctuating pattern reflects the 10 successive local outbreaks within superclusters. Individual model runs show some variation, but the overall pattern is very robust. The chosen time schedule for lifting control means that the last supercluster in the country will lift control 840 days (2 years and 4 months) after the first supercluster. On average individuals in the overall population will experience 432 days (about 14 months) of intensive control. Figure 1B illustrates how the superclusters successively contribute to the overall number of Covid-19 cases over time, averaged over the 8 runs. Figure 1C illustrates the change in the proportion of individuals in the four disease states over time. At the end of the epidemic, 56% of the population has become immune. This is somewhat less than the expected 60% when not accounting for heterogeneity and selection of those with the highest contact rates. This selection process is further illustrated in Figure 1D by the initially diverging average relative contact rates for those susceptible and immune, which eventually become 0.70 and 1.24, respectively; the average relative contact rate of infected and immune cases even touches 1.7 in the first phase of the epidemic. A visualization of the simulated spread of Covid-19 between individuals, clusters, and superclusters is available <u>here</u>. [*URL follows*]

Our proposed strategy goes beyond ‘flattening’ the curve;^3^ with our strategy we ‘tame’ the curve. An important policy-relevant advantage is that instructions to the public will be clear by offering only two regimens: (1) continue the current regulations and recommendations regarding physical distancing and travel restrictions; and (2) permanently return to the normal situation as before Covid-19. This is much more practical than trying to define and achieve a nationwide level of control to not exceed the IC threshold. Also, the strategy to flatten the curve is less efficient because the IC capacity will be optimally used only at the peak of the epidemic (see Supplement 2). A strategy of implementing different intensities of control for different age groups, or implementing alternating periods of control and no control, ^6^ will also be difficult to implement and comes at a relatively high risk of overshooting the threshold. Our strategy will be easier to communicate to the public and thereby likely be more acceptable.

Supplement 3 shows how the results further depend on strategy adjustments and alternative assumptions regarding all model parameters. Basically, the overall duration of the strategy is determined by the chosen number of subdivisions of the country, with a higher number leading to a longer overall duration but lower prevalent case numbers. The required number of superclusters will depend most on *R*_0_ and the chosen maximum number of prevalent Covid-19 cases (dashed horizontal line in Figure 1). A minimum condition for implementing this strategy is that there is sufficiently intensive control (now assumed to reduce contact rates on average to 25% of their original level) to avoid outbreaks in locations that have not yet lifted control. Notably, under this condition, isolation of the supercluster lifting control is not strictly necessary. Also, our strategy can start at any moment, which allows the health care system to prepare for the lift of control in the first supercluster. We further show that it is not strictly necessary that all individuals return to their normal daily activities; however, if many maintain their reduced contact behavior until the end of the strategy, this may trigger a substantially larger last peak. The strategy may be optimized by using an adaptive approach where the next moment of lifting control depends on the state of the outbreak in the preceding supercluster, which would also allow us to counter unexpected outcomes related to uncertainties in Covid-19 transmission. In turn, data resulting from local outbreaks will reduce such uncertainties and help to further improve the strategy and the model.

We realize that the proposed strategy of a phased lift of control still results in many people experiencing a long period of ongoing intensive control, which will undoubtedly cause serious economic and social disruption of (that part of) the country. However, with an ever-growing proportion of the country free of interventions, more people can provide the necessary financial, material and social support to those still experiencing stringent control or a temporary local Covid-19 outbreak. Clearly, the strategy will have a profound impact on individuals and society, and should therefore be considered carefully by various other disciplines (e.g. health systems, ethics, economics) before actual implementation. For instance, we realize that the decision about the order of locations to stop interventions will be extremely complex, leading to political debate and possible societal counteractions. It is conceivable that in the end economic considerations may drive this decision, but in our modelling we have decided to conveniently use a random order, apart from the first (initially most affected) supercluster.

All calculations above can be tailored to country-specific contexts, such as the sizes and composition of administrative units. Also, our strategy will become more efficient when more IC facilities become available, as control can then be lifted simultaneously in multiple areas. In addition, new drugs may reduce the proportion of infected cases requiring IC or the duration of their admission, allowing further shortening of the strategy. We are working on a user-friendly version of the model to support strategy design. As the perfect solution – a vaccine – may well be a matter of years, we conclude that our proposed exit strategy of a phased lift of control should be considered as a way to minimize the Covid-19 disease burden and mitigate the economic and social consequences of prolonged stringent control.

## Data Availability

The source code for the model that was used to simulate data is publicly available (see link).

https://www.gitlab.com/luccoffeng/virsim

## Acknowledgements

We gratefully acknowledge Nico Nagelkerke, Sjoukje Osinga, Epke Le Rutte, Federica Giardina, Wilma Stolk, and Prof. Myriam Hunink for their comments and suggestions. We thank Prof. Diederik Gommers and Prof. Jan Hazelzet for providing quick estimates about IC admissions and availability in the Netherlands.

## Funding

LEC acknowledges funding from the Dutch Research Council (NWO, grant 016.Veni.178.023).

## Conflict of interest

Both authors have no conflict of interest to declare.

## Notes

### Competing Interest Statement

The authors have declared no competing interest.

